# Measuring Disparities in the Impact of COVID-19 on Pediatric Mental Health in Primary Care Settings

**DOI:** 10.1101/2024.08.26.24312603

**Authors:** Cecilia Rogers, Katherine Boguszewski, Angela Gummadi, Mark Conaway, Laura Shaffer, Irène Mathieu

## Abstract

**Objectives:** To examine disparities in mental health diagnosis, depression screening, and depressive symptoms in pediatric primary care settings before and during the COVID-19 pandemic, and to evaluate the use of electronic health records to study temporal trends in pediatric mental and behavioral health (MBH).

**Methods:** This is an IRB-approved, retrospective study of pediatric patients (n=10,866) who visited three primary care sites at an academic medical center before (2017-2019) and during (2020-2022) the COVID-19 pandemic. We used logistic regression to compare rates of diagnoses, depression screening, and depression symptom scores among demographic groups.

**Results:** This study demonstrates an increase in both PHQ-9A screening rates and average scores from 2017-2019 to 2020-2022. There were significant disparities in common mental health diagnoses, including higher rates of psychological distress among lower income and Hispanic patients, both before and during the pandemic, despite lower rates of screening among Hispanic patients. This suggests a need for improved equity in routine MBH screening and additional research to better understand the underlying social determinants that may be driving the greater mental health burden for certain marginalized youth.

This study also highlights the strengths and challenges of utilizing EHR data to characterize disparities in pediatric mental illness. Although the nature of care delivery in an academic medical center clinic and the limitations of the EHR for collecting relevant data present challenges to this measurement, the EHR is nevertheless a promising tool for measuring and tracking pediatric mental health disparities.

## Introduction

The COVID-19 pandemic had significant negative effects on child and adolescent mental and behavioral health, widening the critical gap between patients’ needs and mental health services.^1^ There are multiple reasons for this, including school disruptions in the form of virtual or hybrid education, pandemic-related fear and anxiety, grief and loss, familial social and economic stressors related to illness, decreased income, unemployment, and more.^19, 20^

The pandemic did not affect all demographic groups equally. Children from higher-income and White families were more likely to attend school in person rather than remotely.^2^ In-person education was associated with fewer mental health difficulties, whereas African American and Hispanic older children attending remote school were the most likely to experience emotional difficulties.^2^ Front-line service and low-wage workers have been disproportionately affected by the stressors induced by the pandemic’s disruption, and these stressors have demonstrated negative impact on children’s mental health.^3,4^

The results of these mental health stressors have been observed in both primary care and emergency room settings. In an analysis of results of the Patient Health Questionnaire-9 Modified for Adolescents (PHQ-9A) and the Ask Suicide-Screening Questions (ASQ), administered at 12 primary care practices from June 2019 to October 2020 among patients primarily on Medicaid, over half of patients screened positive for any form of depression, and one-quarter for major depressive disorder (MDD).^5^

Now that children have returned to in-person education and pandemic-related emergency measures have been lifted in most localities, there is a need to better understand and address mental health disparities in the aftermath of the pandemic. To this end, we reviewed records in the electronic health record (EHR) to quantify the burden of mental health symptoms and diagnoses at three primary care clinic sites associated with an academic medical center serving Charlottesville, Virginia and surrounding counties. EHR enables the collection of large amounts of data; however, there are also documented limitations of EHR in quantifying mental health disorders^32^. These limitations include missing mental health-related information and biased documentation of sensitive information.^12^ Other confounding factors that affect the use of EHR include financial, technical, and organizational factors, as the availability of EHR data varies among health systems.^13^

Emerging data illustrate worsening trends in youth mental health in the context of the pandemic. Beyond the physical effects of COVID, social distancing, school closure, and isolation negatively impacted the mental health of children and adolescents.^19, 20^ Outcomes for youth mental health during the pandemic included stress, hopelessness, and risky behavior patterns such as increased alcohol and cannabis use.^19, 21^ Overall increases in anxiety and depressive symptoms also were observed in pediatric populations.^19,20, 21^ Evidence also shows an almost three-fold increase in youth requiring mental stabilization for eating disorders since the start of the pandemic.^22^

This study aims to describe demographic differences in the diagnosis of common mental health disorders, including depression, anxiety, and externalizing behaviors (oppositional defiant disorder, conduct disorder), rates of depression screening and levels of depressive symptomatology, as well as to evaluate the use of EHR for studying pediatric mental health.

Throughout this paper, we will use terminology to refer to racial/ethnic categories consistent with the terminology of our EHR.

## Methods

This study was approved by the University of Virginia Institutional Review Board for Health Sciences Research as exempt. One team member (CR) conducted a literature review to generate a list of relevant diagnoses^14,15^. A general pediatrician (IM) and a child psychologist (LS) then reviewed this list and added to it based on their clinical expertise. For a complete list of diagnoses please see Appendix 1. We were interested in characterizing demographic differences in the prevalence of diagnoses based on race/ethnicity, age, socio-economic status, neighborhood, and sex.

Data for this retrospective chart review were collected using the SlicerDicer tool in Epic. This tool uses different base models to retrieve data from EHRs. In this study, the Visits data model was initially used to extract data. Data were further refined by the addition of other parameters and characteristics central to the research question, as outlined below. Data were then used to compare variables within a baseline population. The timeframe of the study included two cohorts, 1/1/2017-1/1/2019 (inclusive) and 1/1/2020-8/1/2022 (inclusive), to capture data pre-pandemic and during the height of COVID-19. Patient encounters were pulled from three general pediatric clinics affiliated with an academic medical center - one serving primarily the urban population of a small city, one serving a primarily suburban population of the same city, and one serving a rural community approximately 50 minutes outside of the city. Visit type was limited to *Peds Sick Visit or Peds Preventative*. The baseline was also limited to visits with a completed status. Once the study population was refined (N=10,866), prevalence of diagnoses was studied. Five groups of mental health diagnoses (Appendix, Table 1) were added by ICD-10 codes using the EDG Concept tool in SlicerDicer; this tool is a diagnosis grouper used to list related diagnosis codes as the same condition. For example, all ICD-10 codes related to major depressive disorder are grouped under EDG Concept Depression.

Demographics (Appendix Table 2) pulled with each visit included *MRN, Patient Name, Address, City, County, Postal Code, Patient Race, Gender Identity, Legal Sex, Date of Birth, Age in Years, Patient Ethnic Group, Language, and Primary Payer*. During analyses, the data were de-identified by assigning arbitrary “patient study numbers” to each MRN and additional protected health information were then removed from the data set. Multiple encounters for the same patient were identified through use of the patient study numbers. The data were cleaned to combine multiple encounters for the same patient into one row so all diagnoses per patient would be seen together. Each column for diagnoses was then coded using binary variables with 1 indicating the presence of a diagnosis in at least one encounter and 0 indicating an absence of the diagnosis across all encounters analyzed.

PHQ-9A documentation and scores were used to collect data concerning depression screening and symptomatology. The population (N=6,782) for PHQ-9A screenings was limited to 11-18 year olds seen for a Peds Preventive visit, in order to sample patients that should have received a PHQ-9A score per preventive care protocols.

We used chi-squared and logistic regression analyses to determine demographic characteristics associated with diagnoses of interest (led by MC). Rates of screening were also analyzed using chi squared and logistic regression. We compared average PHQ-9A values using 2 sample t-tests and linear regression.

## Results

In the 2017-2019 cohort the patient population was 49% male and 51% female. 1.62% of all males had a mental health diagnosis compared to 2.18% of females. The racial makeup of the study sample was 64% Caucasian, 22% African American, 2% Asian, 4% more than one race, and 8% other. Diagnosis rates in each of these categories were 1.97%, 1.98%, 0.00%, 2.36%, and 1.48% respectively. Non-Hispanic patients made up 86% of the population while Hispanic patients made up 13%. Non-Hispanic patients were diagnosed at a rate of 1.98% while Hispanic patients had a diagnosis rate of 1.41%. Patients were also grouped by insurance group: Medicare/Medicaid, Military, None, Private. Percentage of patients in each insurance group were 42%, 2%, 5%, 50% respectively, and diagnosis rates among insurance groups were 2.16%, 1.90%, 2.68%, 1.61% respectively.

In the 2020-2022 cohort the patient population was 50% male and 50% female. 1.56% of all males had a diagnosis compared to 2.73% of females. By race, the sample was 63% Caucasian, 21% African American, 2% Asian, 4% more than one race reported, and 10% other. The diagnosis rates of the racial categories were 2.09%, 1.99%, 1.06%, 2.71%, 2.85% respectively. Non-Hispanic patients made up 85% of the population while Hispanic patients made up 14%. Non-Hispanic patients were diagnosed at a rate of 1.86% while Hispanic patients had a diagnosis rate of 3.80%. Percentage of patients in each insurance group were 42% Medicare/Medicaid, 2% Military, 5% None, 51% Private. Diagnosis rates among insurance groups were 3.08%, 0.65%, 2.84%, 1.37% respectively. More extensive demographic data can be seen in the Appendix Table 2.

All data on average PHQ-9A scores, screening rates, and rates of diagnoses are portrayed in tables 3-6 in the appendix as well as Figure 1. The prevalence of diagnosis type among demographic groups was not analyzed because the population of patients with a diagnosis was too small to achieve significant results when further disaggregating the population. Therefore, the rate of diagnosis refers to patients with any of the included diagnoses in Table 1 of the Appendix.

**Figure 1.**
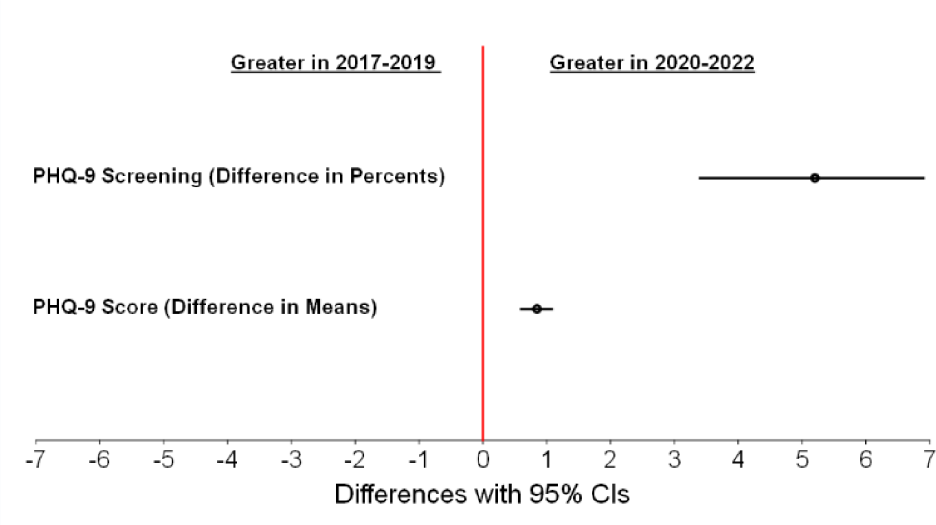
Demonstrates an increase in both PHQ-9A screening rates and average scores from 2017-2019 to 2020-2022.

During 2020-2022, Hispanic patients were more likely to have a mental health diagnosis than non-Hispanic patients (OR 2.09, 95% CI 1.49-2.94). Hispanic patients had a higher rate of mental health diagnoses compared to non-Hispanic patients. However, we also observed that Hispanic youth were less likely to be screened for depression than their non-Hispanic counterparts (OR 0.84, 95% CI 0.71-1.00).

In both cohorts, patients with public (2017-2019: OR 2.60, 95% CI 2.29-2.96; 2020-2022: OR 1.70, 95% CI 1.50-1.96) or military insurance (2017-2019: 4.39, 95% CI 2.65-7.28; 2020-2022: OR 2.62, 95% CI 1.55-4.43) were more likely to be screened than patients with private insurance. Public (2017-2019: OR 1.03, 95% CI 0.62-1.45, p=0.000; 2020-2022: Est. 1.29, 95% CI 0.89-1.68, p= 0.000) and military (2017-2019: Est. 1.55, 95% CI .29-2.82, p=0.016) insurance holders also had higher average PHQ-9A scores. Furthermore, in 2022, patients with Medicaid (OR 2.27, 95% CI 1.65-3.12) or no insurance (OR 2.13, 95% CI 1.65-3.12) had higher prevalence of diagnoses than those with private insurance.

In both cohorts, female patients had higher average PHQ-9A scores (2017-2019: est. 0.85, 95% CI 0.47-1.24, p=0.000; 2020-2022: est. 1.70, 95% CI 1.33-2.07, p=0.000) than male patients. They had higher rates of diagnoses (OR 1.77, 95% CI 1.30-2.41) during 2020-2022 as well. Furthermore, whereas both male and female average PHQ-9A scores increased from 2017-2019 to 2020-2022, females’ scores increased significantly more thanthan males’ (diff 0.85, 95% CI 0.37-1.32, p=0.000).

## Discussion

### Pediatric Mental Health Before and During the Pandemic

The simultaneous increase in overall screening rates and average PHQ-9A scores suggests that both psychological distress and depression screening rates increased during the COVID-19 pandemic. It is possible that the lower average PHQ-9A scores pre-pandemic reflect an underestimation of distress confounded by the lower incidence of depression screening. However, a more likely explanation of the data in keeping with clinical experience is that psychological distress increased during the pandemic inherently increased psychological distress and providers began screening more patients more frequently in response. Routine screening is imperative for early detection and intervention as well as for tracking trends in symptomatology over time and in different populations.

Using insurance as a proxy for socio-economic status, our findings suggest a disproportionate burden of mental health disorders on populations with lower SES before and during the COVID-19 pandemic. This insight demonstrates the necessity of facilitating access to care for low-income families who were more vulnerable to the adverse effects of the COVID-19 pandemic. The data also reveal concerning disparities for Hispanic patients at the height of the pandemic. These youth had higher prevalence of mental health diagnosis and lower rates of depression screening than their non-Hispanic peers. This discrepancy is worrisome in that it clearly indicates a greater burden of distress in Hispanic youth, given diagnosis rates alone that could well indicate an underestimate of psychological distress and even greater disparity that went undetected because of the lower screening rate. The impact of language barriers for this population in screening rates, scores, and rates of diagnoses should be considered and further studied.

The higher average PHQ-9A scores and rates of mental health diagnoses for female patients may indicate the bias of mental health screening towards internalizing symptoms, which are more prevalent in girls as compared to boys. This suggests a need for more comprehensive mental health screening that includes assessments of externalizing as well as internalizing symptoms.

The relatively low rates of mental health diagnoses in our sample make it difficult to compare differences across race/ethnicity categories, despite the large overall sample size. We were unable to draw conclusions about geographic disparities for the same reason. It could also represent a bias in which patients who were doing relatively better and with fewer stressors were more likely to come into clinic during the pandemic. It may also be due to heterogeneity of documentation and coding practices, as well as the limitations of the EGD concept category in SlicerDicer for capturing all possible diagnostic codes providers may use. Furthermore, this finding underscores the importance of more consistent screening and documentation, as well as the need for additional multi-site trials that would increase power.

### Strengths and Challenges of the EHR

#### Strengths

Mental and behavioral health challenges are common concerns discussed in both sick and well visits to pediatric primary care providers (PCP). Many symptoms that are discussed and receive diagnostic codes from a PCP may not meet criteria or escalate to become a diagnosable mental illness, such as major depressive disorder. However their discussion at a visit may represent significant parental concern, child distress, and/or interference with daily functioning. Therefore the inclusion of nonspecific, symptom-based diagnostic codes may provide a more accurate survey of a pediatric population’s mental wellness at a given point in time. We found that including these codes allowed us to capture a fuller spectrum of psychological distress in our population.

The necessity of diagnostic codes for billing means that even absent a formal diagnosis of a mental illness, data about MBH concerns often are readily available in and extractable from the EHR. Furthermore, MBH screening is a regular aspect of care delivered by many PCPs. In the case of the study clinics, PHQ-9A depression screening is a part of routine care protocols for well child check visits for all children aged 11 years and older, and also are administered on an as-needed basis per the PCP’s judgment during other visits. The PHQ-9A scores represent an additional source of information about pediatric mental well-being in the form of a continuous and near-universal variable in the EHR.

#### Challenges

The first challenge is that of representation. EHR data inherently cannot be used to address clinical concerns for those patients who do not come into the clinic. This is particularly significant for MBH, since children whose parents do not recognize they are struggling or who are not able to bring them into the clinic due to work constraints, transportation issues, or other barriers may not receive the appropriate treatment or referrals necessary. This suggests that quantitative studies relying solely on EHR data may underestimate the degree of disparities in MBH concerns within a given population.

The heterogeneity of our clinical settings posed a number of challenges. The high number of providers, including trainees at various stages, results in inconsistencies in documentation. Pertinent information potentially may be found in multiple areas of a patient’s chart, depending on the provider’s personal documentation practices. For instance, some providers document the results of the PHQ-9A in the screening tab designed for this purpose. The PHQ-9A score can also be found in multiple flowsheets. Others type the results as free text in the body of the encounter note. Additionally, symptoms that a parent or patient reports might be charted in a variety of ways, depending on the provider. For instance, after a discussion about a child’s new-onset aggression, some providers may select a code such as “Behavior problem in pediatric patient” while others may document the family’s concerns as free text in the note without selecting a corresponding diagnostic code.

Furthermore, patient-entered demographic documentation such as race and ethnicity is not always available. Some patients choose not to provide this information, and in some cases medical assistants determine and enter this information, with or without input from the patient. There is no ability to determine who enters such data in our EHR. This raised questions of data validity, since information entered by medical assistants may reflect the assistants’ assumptions and not patients’ own self-identification. This is a well-known limitation of data available on patient race and ethnicity in the EHR^34^ (Kader and Chebli). Similarly, little information about socio-economic status (SES) is routinely captured in the EHR. We used insurance type as a proxy for SES, since this information is universally available and generally accurate; however there are limitations to using insurance type as an SES proxy^33^. While all three of our clinic sites routinely screen for social needs such as food and housing insecurity, these data are collected on paper forms and used primarily for connecting patients with resources during clinical encounters, but are not routinely captured in the EHR.

There were also particular issues with the SlicerDicer tool in Epic, which was used as the primary tool for data extraction. Lack of a comprehensive SlicerDicer dictionary generated a lot of confusion around where to find and how to interpret data, which resulted in months of trial and error to pull usable data. The Epic Learning Library has severely limited detail on SlicerDicer. There is an overview of the general use of SlicerDicer, but no specifics on the variables and criteria for potential analysis. For instance, there was a lack of clarity around the exact location in a patient’s chart from which SlicerDicer was pulling ICD-10 codes (whether from progress notes, orders, or referrals, etc.).

Given the public health significance of the pediatric mental health crisis, it is critical to develop methods for accessing large-scale clinical data. One research team at Duke University developed a datamart that provides institutional researchers with more easily accessible and usable data drawn from the EHR^35^ (Hurst et al). The adaptation of a similar platform at other institutions is a promising solution to the challenges outlined in this paper.

## Limitations

Although this study includes a relatively large sample size of patients across three very different clinical settings, all clinics are housed within the same academic medical system, which limits geographic generalizability. Furthermore, while the PHQ-9A is an important and commonly used mental health screening tool, it does not capture other common mental health concerns such as anxiety symptoms or disordered eating. We chose to focus on PHQ-9A screening because it is commonly used across multiple health center contexts and results data is more readily available in our EHR than for any other screening tool. However, the lack of inclusion of other screening instruments limits the types of disparities we could measure in this study. An even larger sample size, ideally across multiple institutions, could provide more information on disparities in less frequently screened symptoms and conditions.

## Conclusion

In this study we document significant disparities in common mental health diagnoses, including higher rates of depressive symptoms among lower income patients both before and during the pandemic, and higher rates of depressive symptoms among Hispanic patients despite lower rates of screening among this population. This suggests a need for improved equity in routine MBH screening and additional research to better understand the underlying social determinants that may be driving these disparities.

This study also highlights the strengths and challenges of utilizing EHR data to characterize disparities in pediatric mental illness, which is a growing crisis in the United States. While many have called for greater attention to pediatric mental health in primary care settings, an important first step is to measure the scope of the problem and identify sub-populations at greater risk of mental illness. Although the nature of care delivery in an academic medical center clinic and the limitations of the EHR for collecting relevant data present challenges to this measurement, the EHR is nevertheless a promising tool for measuring and tracking pediatric mental health disparities. This study and others like it may provide important information for improving the EHR for the purposes of tracking population-level mental health and institutional quality improvement.

## Data Availability

Research data are not shared.

## Data availability statement

Research data are not shared.

## Ethics approval statement

This research was approved by the University of Virginia Institutional Review Board for Health Sciences Research as an exempt study and has granted a waiver of HIPAA authorization under 45CFR 164.512(i)(2) via expedited review procedures for the main study.

## Patient consent statement

N/A

## Permission to reproduce material from other sources

N/A

