## Appendix for "Measuring Disparities in the Impact of COVID-19 on Pediatric Mental Health in Primary Care Settings"

**Table 1** List of ICD-10 codes and corresponding diagnosis

| Diagnosis | ICD-10 Code |
| --- | --- |
| Depression | EDG Concept Depression, group of 1785 codes for depression disorders |
| Anxiety | EDG Concept Anxiety, group of 1312 codes for anxiety disorders |
| Eating Disorder | EDG Concept Eating Disorder, Group of 260 codes for eating disorders |
| Oppositional Disorder | 181 codes for ICD-10-CM: F91* |
| Conduct Disorders, unspecified | 46 codes for ICD-10-CM: F91.9 |

**Table 2** Demographics*

| Characteristic | Category | 2017-2019 | | | 2020-2022 | | |
|---|---|---|---|---|---|---|---|
| | | Number In category | Number with any dx | % with any dx | | Number with any dx | % with any dx |
| Sex | Male | 3763 | 61 | 1.62% | 4179 | 65 | 1.56% |
| | Female | 3860 | 84 | 2.18% | 4173 | 114 | 2.73% |
| Age at initial | 5-9 | 3391 | 26 | 0.77% | 3528 | 33 | 0.94% |
| | 10-14 | 2835 | 58 | 2.05% | 3178 | 73 | 2.30% |
| | 15-18 | 1397 | 61 | 4.37% | 1646 | 73 | 4.43% |
| Race | Caucasian | 4880 | 96 | 1.97% | 5221 | 109 | 2.09% |
| | African American | 1669 | 33 | 1.98% | 1762 | 35 | 1.99% |
| | Asian | 164 | 0 | 0.00% | 188 | 2 | 1.06% |
| | More than 1 | 296 | 7 | 2.36% | 369 | 10 | 2.71% |
| | Other | 607 | 9 | 1.48% | 807 | 23 | 2.85% |
| Ethnicity | Non-Hispanic | 6610 | 131 | 1.98% | 7110 | 132 | 1.86% |
| | Hispanic | 992 | 14 | 1.41% | 1210 | 46 | 3.80% |
| Insurance | Medicare/Medicaid | 3195 | 69 | 2.16% | 3471 | 107 | 3.08% |
| | Military | 158 | 3 | 1.90% | 155 | 1 | 0.65% |
| | None | 410 | 11 | 2.68% | 423 | 12 | 2.84% |
| | Private | 3848 | 62 | 1.61% | 4295 | 59 | 1.37% |
| Legal Sex/Gender Identity | F-Match | 3823 | 80 | 2.09% | 4139 | 112 | 2.71% |
| | F-Not Match | 37 | 4 | 10.81% | 34 | 2 | 5.88% |
| | Male Match | 3747 | 61 | 1.63% | 4165 | 65 | 1.56% |
| | M-Not Match | 16 | 0 | 0.00% | 14 | 0 | 0.00% |
| English Lang | No | 562 | 6 | 1.07% | 789 | 25 | 3.17% |
| | Yes | 7061 | 139 | 1.97% | 7563 | 154 | 2.04% |
| Ethnicity/ Language | NonHisp, English | 6599 | 130 | 1.97% | 7092 | 132 | 1.86% |
| | Hisp, English | 441 | 9 | 2.04% | 439 | 21 | 4.78% |
| | Hisp, Spanish | 551 | 5 | 0.91% | 771 | 25 | 3.24% |

*2017-2019 N=7681; 2020-2022 N=8345

**Table 3** Difference in average PHQ-9A scores from reference category, within cohorts

| Variable | Category | 2017-2019 | | | | 2020-2022 | | | |
|---|---|---|---|---|---|---|---|---|---|
| | | est | LL | UL | P | est | LL | UL | P |
| Sex | Male | ref | | | | | | | |
| | Female | 0.85 | 0.47 | 1.24 | 0.000 | 1.70 | 1.33 | 2.07 | 0.000 |
| Age | 11-14 | 0.39 | -0.02 | 0.79 | 0.061 | 0.20 | -0.18 | 0.58 | 0.307 |
| | 15-17 | ref | | | | | | | |
| Race | Caucasian | ref | | | | | | | |
| | African American | 0.21 | -0.24 | 0.65 | 0.357 | 0.52 | 0.07 | 0.96 | 0.023 |
| | Asian | -1.01 | -2.77 | 0.75 | 0.259 | -1.51 | -3.16 | 0.14 | 0.072 |
| | More than 1 | 0.94 | -0.14 | 2.02 | 0.087 | 1.58 | 0.53 | 2.64 | 0.003 |
| | Other | -0.04 | -0.78 | 0.69 | 0.905 | 0.13 | -0.55 | 0.81 | 0.710 |
| Ethnicity | Non-Hispanic | ref | | | | | | | |
| | Hispanic | 0.07 | -0.50 | 0.64 | 0.804 | 0.20 | -0.33 | 0.73 | 0.465 |
| Insurance | Medicare/Medicaid | 1.03 | 0.62 | 1.45 | 0.000 | 1.29 | 0.89 | 1.68 | 0.000 |
| | Military | 1.55 | 0.29 | 2.82 | 0.016 | 1.14 | -0.11 | 2.38 | 0.073 |
| | None | 0.31 | -0.38 | 0.99 | 0.381 | 0.44 | -0.42 | 1.29 | 0.315 |
| | Private | ref | | | | | | | |
| Legal Sex/Gender Identity | Match | -2.67 | -4.34 | -1.00 | 0.002 | -3.97 | -6.02 | -1.92 | 0.000 |
| | Not Match | ref | | | | | | | |
| English Lang | No | ref | | | | | | | |
| | Yes | 1.08 | 0.14 | 2.02 | 0.024 | 0.13 | -0.55 | 0.80 | 0.717 |
| Ethnicty/ Language | NonHisp,Eng | ref | | | | | | | |
| | Hisp, Eng | 0.61 | -0.07 | 1.29 | 0.081 | 0.60 | -0.16 | 1.36 | 0.122 |
| | Hisp, Spanish | -0.97 | -1.92 | -0.02 | 0.045 | -0.13 | -0.81 | 0.56 | 0.714 |

**Table 4** Estimation of Odds Ratios for Screening, within cohorts

| Variable | Category | 2017-2019 | | | | 2020-2022 | | | |
|---|---|---|---|---|---|---|---|---|---|
| | | OR | LL | UL | P | OR | LL | UL | P |
| Sex | Male | ref | | | | | | | |
| | Female | 1.15 | 1.02 | 1.29 | 0.019 | 1.03 | 0.91 | 1.17 | 0.650 |
| Age in 2017 cohort | 11-14 | 0.76 | 0.67 | 0.86 | 0.000 | 0.73 | 0.65 | 0.83 | 0.000 |
| | 15-17 | ref | | | | ref | | | |
| Race | Caucasian | ref | | | | | | | |
| | African American | 1.60 | 1.39 | 1.84 | 0.000 | 1.20 | 1.02 | 1.40 | 0.025 |
| | Asian | 0.50 | 0.32 | 0.78 | 0.002 | 0.56 | 0.36 | 0.89 | 0.014 |
| | More than 1 | 2.67 | 1.78 | 4.00 | 0.000 | 1.36 | 0.91 | 2.04 | 0.130 |
| | Other | 0.97 | 0.78 | 1.20 | 0.782 | 0.72 | 0.58 | 0.89 | 0.002 |
| Ethnicity | Non-Hispanic | ref | | | | | | | |
| | Hispanic | 1.04 | 0.88 | 1.24 | 0.631 | 0.84 | 0.71 | 1.00 | 0.049 |
| Insurance | Medicare/Medicaid | 2.60 | 2.29 | 2.96 | 0.000 | 1.71 | 1.50 | 1.96 | 0.000 |
| | Military | 4.39 | 2.65 | 7.28 | 0.000 | 2.62 | 1.55 | 4.43 | 0.000 |
| | None | 1.68 | 1.37 | 2.06 | 0.000 | 0.92 | 0.70 | 1.20 | 0.519 |
| | Private | ref | | | | | | | |
| Legal Sex/Gender Identity | Match | 0.99 | 0.61 | 1.62 | 0.976 | 1.28 | 0.64 | 2.54 | 0.486 |
| | Not Match | ref | | | | | | | |
| English Lang | No | ref | | | | | | | |
| | Yes | 1.69 | 1.31 | 2.19 | 0.000 | 1.16 | 0.94 | 1.43 | 0.169 |
| Ethnicty/ Language | NonHisp,Eng | ref | | | | | | | |
| | Hisp, Eng | 1.56 | 1.25 | 1.95 | 0.000 | 0.83 | 0.64 | 1.07 | 0.155 |
| | Hisp, Spanish | 0.62 | 0.47 | 0.80 | 0.000 | 0.85 | 0.69 | 1.05 | 0.136 |

**Table 5** Estimation of Odds Ratios for Diagnoses, within Cohorts

| Characteristic | Category | 2017-2019 | | | 2020-2022 | | |
|---|---|---|---|---|---|---|---|
| | | Number In category | Number with any dx | % with any dx | | Number with any dx | % with any dx |
| Sex | Male | 3763 | 61 | 1.62% | 4179 | 65 | 1.56% |
| | Female | 3860 | 84 | 2.18% | 4173 | 114 | 2.73% |
| Age at initial | 5-9 | 3391 | 26 | 0.77% | 3528 | 33 | 0.94% |
| | 10-14 | 2835 | 58 | 2.05% | 3178 | 73 | 2.30% |
| | 15-18 | 1397 | 61 | 4.37% | 1646 | 73 | 4.43% |
| Race | Caucasian | 4880 | 96 | 1.97% | 5221 | 109 | 2.09% |
| | African American | 1669 | 33 | 1.98% | 1762 | 35 | 1.99% |
| | Asian | 164 | 0 | 0.00% | 188 | 2 | 1.06% |
| | More than 1 | 296 | 7 | 2.36% | 369 | 10 | 2.71% |
| | Other | 607 | 9 | 1.48% | 807 | 23 | 2.85% |
| Ethnicity | Non-Hispanic | 6610 | 131 | 1.98% | 7110 | 132 | 1.86% |
| | Hispanic | 992 | 14 | 1.41% | 1210 | 46 | 3.80% |
| Insurance | Medicare/Medicaid | 3195 | 69 | 2.16% | 3471 | 107 | 3.08% |
| | Military | 158 | 3 | 1.90% | 155 | 1 | 0.65% |
| | None | 410 | 11 | 2.68% | 423 | 12 | 2.84% |
| | Private | 3848 | 62 | 1.61% | 4295 | 59 | 1.37% |
| Legal Sex/Gender Identity | F-Match | 3823 | 80 | 2.09% | 4139 | 112 | 2.71% |
| | F-Not Match | 37 | 4 | 10.81% | 34 | 2 | 5.88% |
| | Male Match | 3747 | 61 | 1.63% | 4165 | 65 | 1.56% |
| | M-Not Match | 16 | 0 | 0.00% | 14 | 0 | 0.00% |
| English Lang | No | 562 | 6 | 1.07% | 789 | 25 | 3.17% |
| | Yes | 7061 | 139 | 1.97% | 7563 | 154 | 2.04% |
| Ethnicity/ Language | NonHisp, English | 6599 | 130 | 1.97% | 7092 | 132 | 1.86% |
| | Hisp, English | 441 | 9 | 2.04% | 439 | 21 | 4.78% |
| | Hisp, Spanish | 551 | 5 | 0.91% | 771 | 25 | 3.24% |

**Table 6** Difference in difference of average PHQ-9A scores between cohorts

| | | Diff | Lower 95% | Upper 95% | P |
|---|---|---|---|---|---|
| Sex | Female vs Male | 0.85 | 0.37 | 1.32 | 0.000 |
| Age | Age 11-14 vs Age 15-18 | 0.19 | -0.34 | 0.72 | 0.483 |
| Race | AfrAmer vs Cauc | 0.31 | -0.25 | 0.87 | 0.279 |
| | Asian vs Cauc | -0.50 | -2.74 | 1.74 | 0.663 |
| | More than 1 vs Cauc | 0.64 | -0.68 | 1.96 | 0.340 |
| | Other vs Cauc | 0.17 | -0.74 | 1.09 | 0.709 |
| Ethnicity | Hispanic vs NonHispanic | 0.13 | -0.58 | 0.83 | 0.728 |
| Insurance | Medicare/Caid vs Private | 0.25 | -0.26 | 0.76 | 0.332 |
| | Military vs Private | -0.42 | -2.00 | 1.17 | 0.605 |
| | Non vs Private | 0.13 | -0.88 | 1.15 | 0.800 |
| Legal vs Identity | Match vs Not Match | -1.30 | -3.65 | 1.04 | 0.275 |
| Language | Spanish vs English | 0.96 | -0.11 | 2.03 | 0.080 |
| Eth/Lang | Hisp/English vs NonHisp,Eng | -0.01 | -0.92 | 0.90 | 0.986 |
| | Hisp,Span vs NonHisp,Eng | 0.84 | -0.24 | 1.92 | 0.129 |
